# Preparation for cardiac procedures: identifying gaps between outpatients’ views and experiences of patient-centred care

**DOI:** 10.1101/2023.02.22.23286327

**Authors:** Kristy Fakes, Trent Williams, Nicholas Collins, Andrew Boyle, Aaron L Sverdlov, Allison Boyes, Rob Sanson-Fisher

**Author notes:** Corresponding author: Dr Kristy Fakes^1,2^, Health Behaviour Research Collaborative, School of Medicine & Public Health University of Newcastle, University Dr, Callaghan NSW 2308 AUSTRALIA.

## Abstract

**Background:** To examine the delivery of patient-centred care and identify any gaps in care perceived as essential by patients; this study examined outpatients’: 1) views on what characterises essential care and 2) experiences of care received, in relation to cardiac catheterisation and subsequent cardiovascular procedures.

**Methods:** A cross-sectional descriptive study was undertaken. Surveys were posted to outpatients who had undergone elective cardiac catheterisation in the prior six months at an Australian tertiary public hospital. Participants completed a 65-item survey to determine: a) aspects of care they perceive as essential for a healthcare team to provide to patients receiving care for a cardiac condition (Important Care Survey); or b) their actual care received (Actual Care Survey). Numbers and percentages were used to calculate the most frequently identified essential care items by patients; and the experiences of care received. Items rated as either ‘Essential’ or ‘Very important’ by at least 80% of participants were determined, reflecting patient endorsement of the importance of the component of care. A gap in patient-centred care was identified as being any item that was endorsed as essential/very important by 80% or more of participants but reported as received by less than 80% of participants.

**Results:** Of 582 eligible patients, 264 (45%) returned a completed survey. 43/65 items were endorsed by over 80% of participants as essential/very important. Of those, for 22 items, less than 80% of respondents reported the care as received. Gaps were identified in relation to GP consultation (3 items), preparation (4 items), having the procedure (2 items), follow-up care (1 item), subsequent decision making for treatment (4 items), prognosis (6 items) and post-treatment follow-up (1 item).

**Conclusions:** Areas were identified where actual care fell short of patients’ perceptions of essential care, particularly general practitioner involvement, the referral process and information on patient prognosis.

## INTRODUCTION

Cardiac catheterisation is a widely performed procedure throughout the world, and whilst typically a relatively safe and well-tolerated procedure with a low complication rate, the potential impacts on morbidity and mortality can be significant.(1) Patient-centred care is an essential, overarching component of quality health care.(2) It is defined as ‘care that is respectful of and responsive to individual patient preferences, needs, and values and ensuring that patient values guide all clinical decisions’.(2) In the context of patient-centred care, significant benefits to patients’ outcomes and experiences of care as well as benefits to the healthcare system, such as clinical quality have been reported.(3, 4)

Measurement of patient-reported outcomes and care experiences play a key role in the transformation towards patient-centred health systems.(5, 6) For example, Patient-Reported Outcome Measures (PROMs) are currently used to obtain patients’ views on health-related quality of life and symptoms.(7-9) In comparison, Patient-Reported Experience Measures (PREMs) are those that obtain reports of what actually occurred during a care event,(10) i.e. the occurrence of concrete and specific components of care, rather than patients’ evaluation of what occurred.(11)

Few studies, however, have examined patient experiences, or the quality of patient-centred care provided to patients who have undergone cardiac catheterisation, and indeed patient-reported experiences of cardiovascular procedures more broadly. A recent review of opportunities for improving patient-reported experiences for cardiovascular disease(6) identified the need for further research into measurement tools. A separate review(12) examined quality improvement frameworks within cardiac catheterisation laboratories.

However, it focussed on clinical outcomes and patient safety, like most other work in relation to cardiac procedures, rather than the improvement of patient experiences and the provision of patient-centred care. Recent consensus statements, including those commissioned by the Society for Cardiovascular Angiography and Interventions (SCAI): *SCAI expert consensus update on best practices in the cardiac catheterization laboratory(13)* include principles for patient experience optimization including patient surveys. However, it is also acknowledged in those guidelines that new surveys would need to be developed to examine patient experiences of the cardiac catheterisation laboratory.

An important component that should be examined is the provision of information. Patient education is essential for informed consent, and to ensure patients are well equipped to take an active role in their preparation and recovery from medical procedures including cardiac catheterisation. However, a previous review found few measures of patient preparation for medical interventions exist.(14) To improve patient outcomes, we need to identify any evidence-practice gaps i.e., a difference between components of care considered essential based on evidence-based guidelines and importance to patients, and the actual care provided to patients. Healthcare providers need access to patients’ views about care and any patient-perceived gaps in care and/or information provision across a broad range of components and phases of care to prioritise healthcare setting and system-level quality improvement initiatives which can enhance the delivery of integrated, patient-centred care.

As such, the need for further research and a new measure to examine patient experiences of cardiac catheterisation was identified. To identify any gaps in patient-centred care, this study specifically examined outpatients’: 1) views on what is characterised as essential or important care and 2) experiences of care in relation to cardiac catheterisation and other subsequent cardiovascular procedures.

## METHODS

### Study type

Cross-sectional descriptive study.

### Population and setting

Participants were recruited from the Cardiology Department, Cardiac Catheterisation Laboratory at a major public hospital in regional Australia which services a population of approximately one million people.

### Recruitment and selection of participants

Cardiology Department staff identified eligible patients utilising hospital procedural lists. People considered eligible were individuals aged 18 years or older who had undergone an elective outpatient cardiac catheterisation in the Cardiac Catheterisation Laboratory at the participating site within the last six months.

All eligible outpatients were sent a recruitment pack via post by staff from the Cardiology Department containing an invitation to participate, information statement, pen-and-paper survey and reply-paid envelope for survey return to the researchers. Reminder packs were sent to all non-responders two weeks later.

### Data Collection

#### Measures

##### Quality of care

To reduce participant burden, patients were either asked for their views about:

a. What characterises essential or important care (Important Care Survey), see below; or
b. Their actual care received (Actual Care Survey), in relation to the same aspects included in the Important Care Survey.

##### Important Care Survey

The items describe components of care along the patient pathway (initial GP consultation and referral process (4 items), preparation for the procedure (11 items), having the procedure (7 items), recovery (4 items), follow-up care (5 items), subsequent decision making for treatment (11 items), prognosis (6 items) and post-treatment follow-up (17 item)).

##### Item development, and face and content validation

The published literature was assessed to identify the needs, concerns, and issues facing patients undergoing cardiac catheterisation. Items of existing instruments were reviewed and considered for inclusion, including instruments assessing patient preparation(15) and life expectancy;(16) as well as the Institute of Medicine’s dimensions of patient-centred care;(2) and supportive and psychosocial care guidelines.(17, 18) Item development was also informed by qualitative interviews undertaken as part of development of a generic instrument that examined preparedness for medical interventions.(15) As reported elsewhere,(15) this included 33 patients undergoing medical imaging procedures including angiography.

The identified items were expanded to specifically address all phases across the cardiac catheterisation care pathway, from the patient’s consultation with their general practitioner (GP) and referral to a cardiologist, to post-treatment follow-up care. To further confirm face validity (the degree to which items are an adequate reflection of the construct to be measured) and content validity (i.e. the degree to which the content of the measure is an adequate reflection of the construct to be measured)(19), the draft items were reviewed by a multidisciplinary team of health behaviour scientists (n = 4), clinicians (n = 5) and consumer representatives (n = 2).

To examine patient endorsement of each component of care/item, participants were asked to indicate how important the criterion was if the best care possible was to be provided to patients. Participants were asked to respond to the 65 statements by choosing one of the following response options: “Essential”, “Very important”, “Moderately important”, “Somewhat important”, or “Not important”. Thirty-one items were about the initial cardiac catheterisation, whilst the remaining 34 items were about any subsequent cardiac-related procedures the patient had, including angioplasty on same or different day; coronary artery bypass grafting; heart valve repair or replacement; pacemaker insertion; and cardiac defibrillator implant.

##### Actual Care Survey

For each of the same components of care included in the Important Care Survey, participants were asked about their receipt of that care or information. E.g. “*Did your [healthcare team] give you information about…?*” Participants responded to 65 statements by choosing “Yes” or “No” to indicate if the aspect of care was received. The yes/ no response format was chosen instead of a Likert scale rating as it enables the participant to report an actual occurrence of care, rather than their perceptions or ratings of the experience.

#### Sociodemographic characteristics

Patient demographic and medical characteristics: age; gender; education; marital status; employment status; living arrangements; travel time to hospital were also assessed in each survey.

#### Sample size and statistical analyses

##### Sample size

Approximately 1200 adult outpatient cardiac catheterisation and percutaneous coronary interventions are performed per year at the participating hospital. Based on a conservative power calculation, a total sample size of 200 participants (100 per survey) was estimated to enable the proportion of patients reporting particular experiences of care with 95% confidence intervals within ±6.9%.

### Statistical analysis

Characteristics of the participants were compared using means, standard deviations, medians and interquartile intervals for continuous variables and by percentages for categorical variables. Frequencies and percentages were used to determine the most frequently identified essential criteria for care for patients; and actual care received. To identify any gaps in patient-centred care, items rated as either ‘Essential’ or ‘Very important’ by at least 80% of participants were determined, reflecting patient endorsement of the importance of the component of care. A cut point of 80% was chosen, being a standard approach for measuring consensus.(20) A gap in care was then identified as being an item that was endorsed as essential/very important by 80% or more of participants, but reported as received by less than 80% of participants.

## RESULTS

### Sample

Overall, out of 582 eligible patients approached for this study, 264 (45%) consented to take part and returned a completed survey. Of those, 131 patients completed the Actual Care Survey and 133 patients completed the Important Care Survey. Patients’ demographic characteristics are reported in Table 1. Participants were aged 40–89 years and underwent their procedure for a variety of underlying cardiac conditions.

**Table 1.**
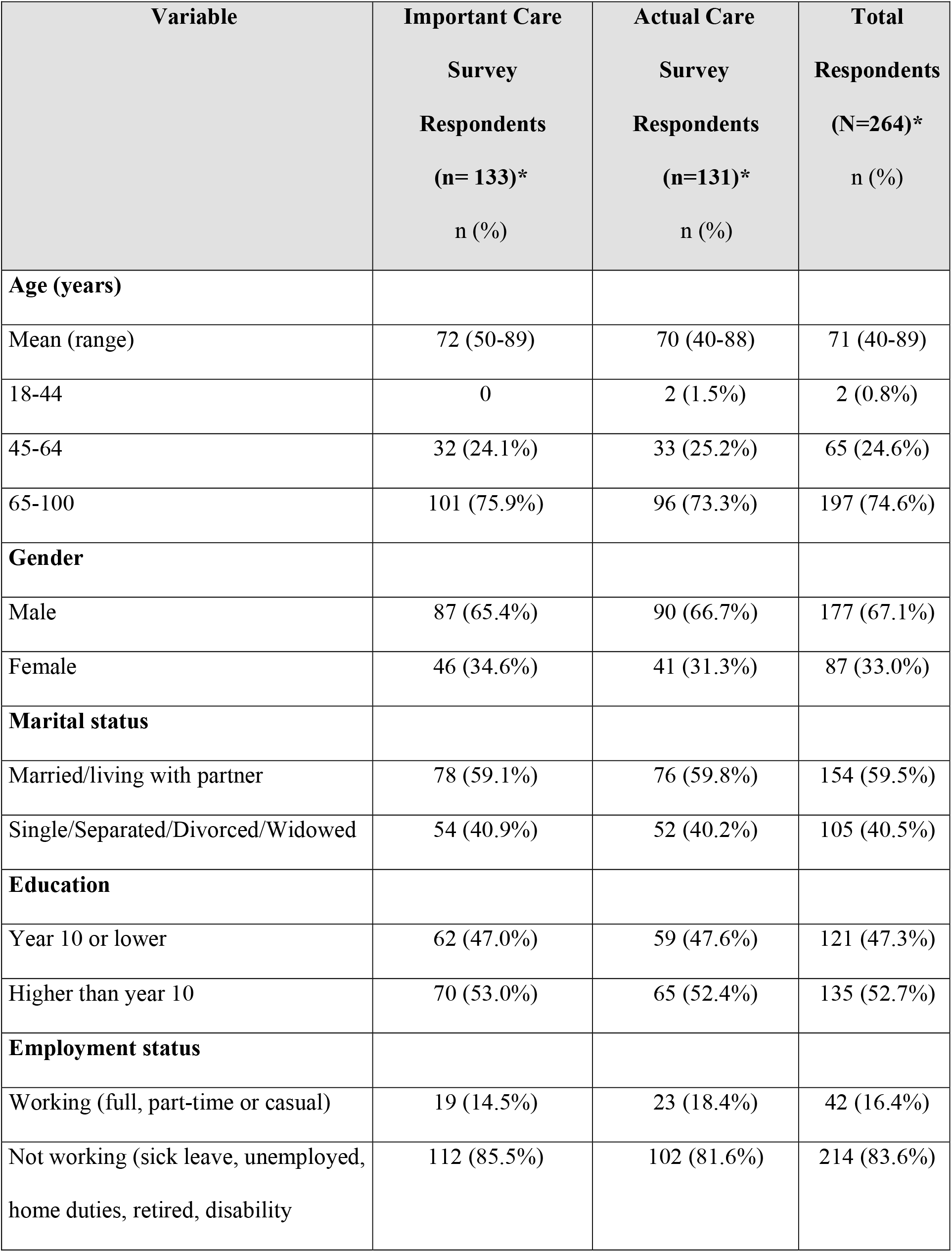
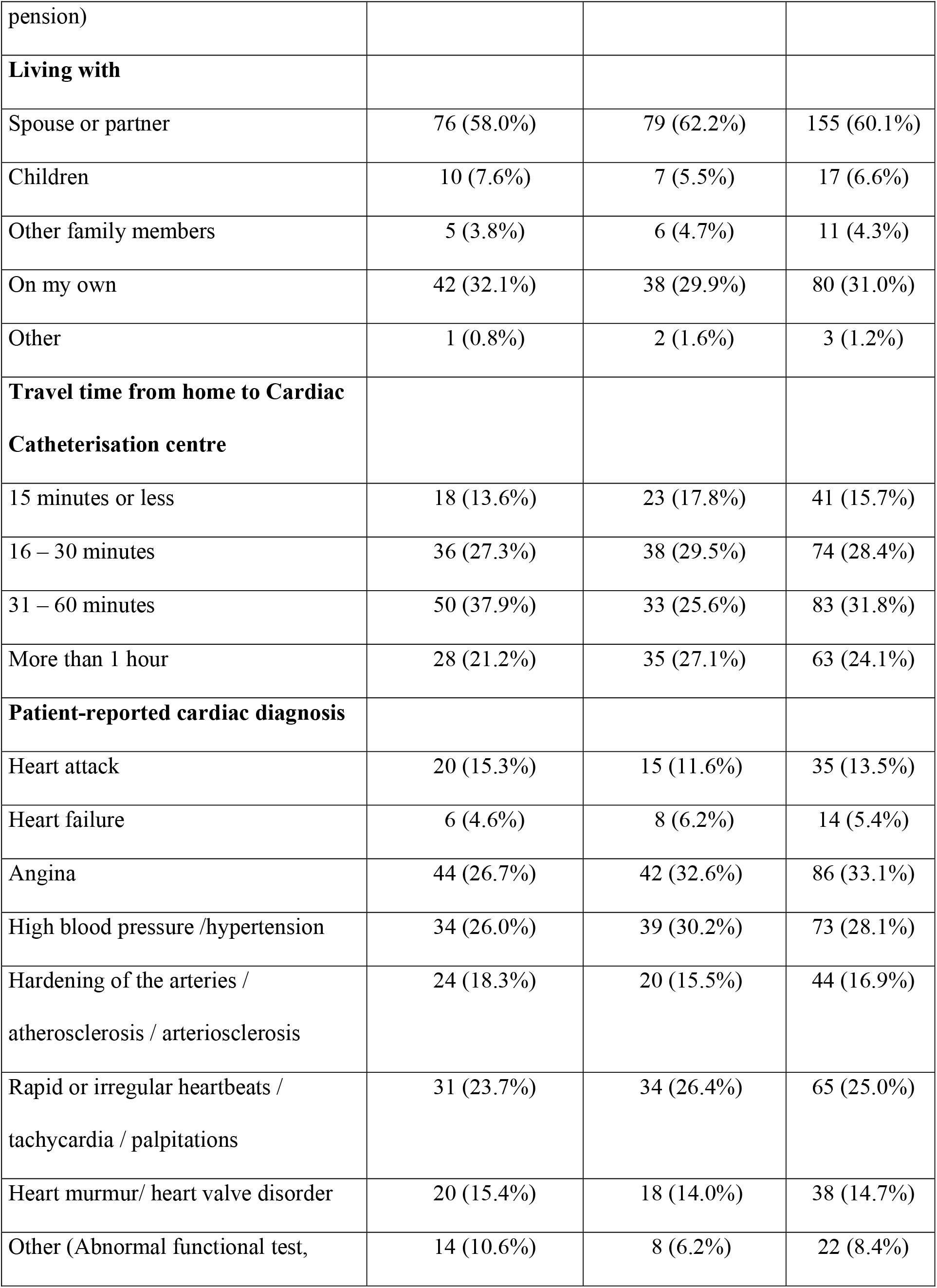
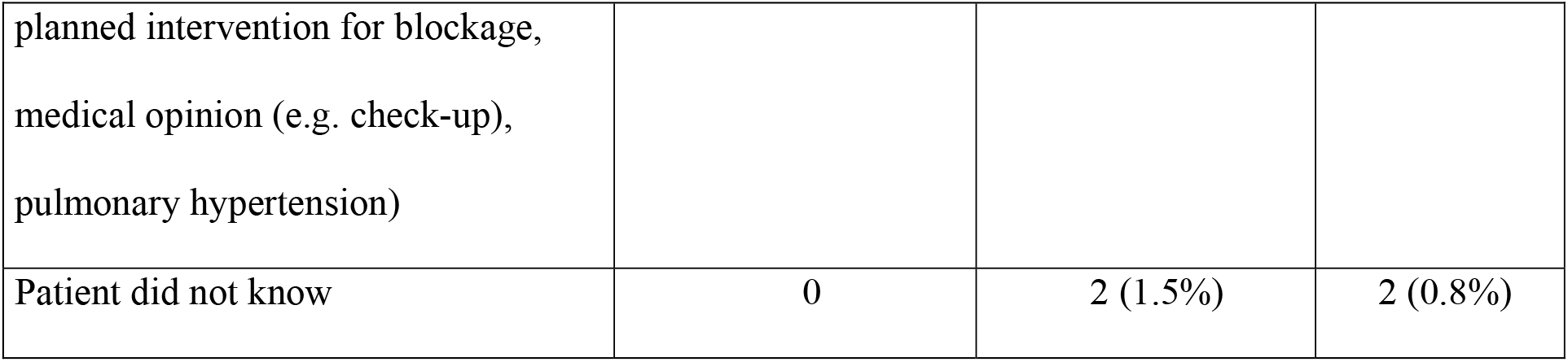
Demographic and disease characteristics of study sample (n=264)

### Patients’ ratings of importance and whether components of care were received

As shown in Table 2, participants completed 31 items identifying either: i) aspects of care that patients perceive are important or essential for a healthcare team to provide in order to best support patients receiving care for a suspected or confirmed heart condition; or ii) their actual care received in relation to the: GP consultation and referral process (4 items); preparation for the procedure (11 items); having the procedure (7 items); recovery (4 items) and follow-up care (5 items).

**Table 2.**
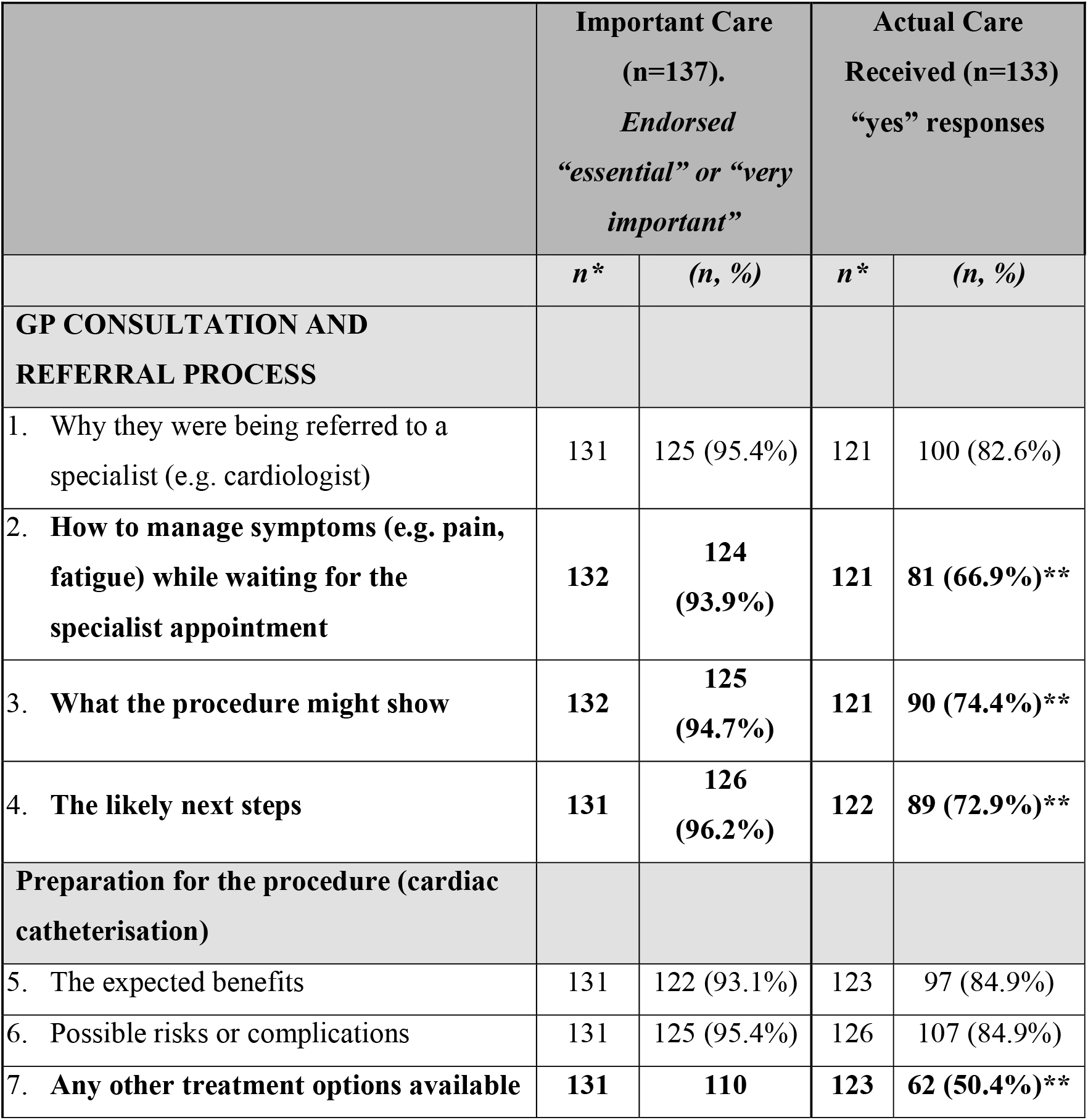
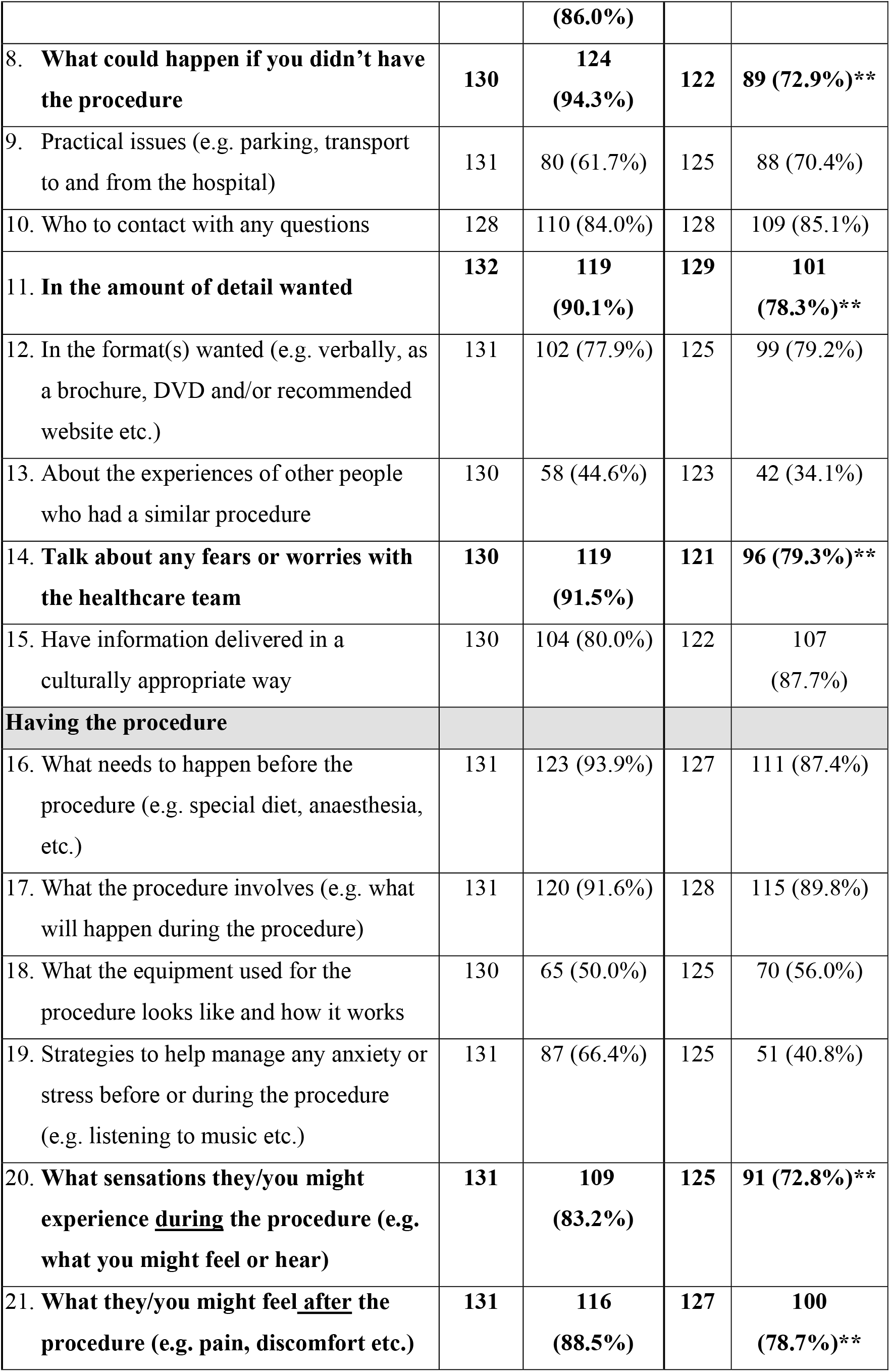
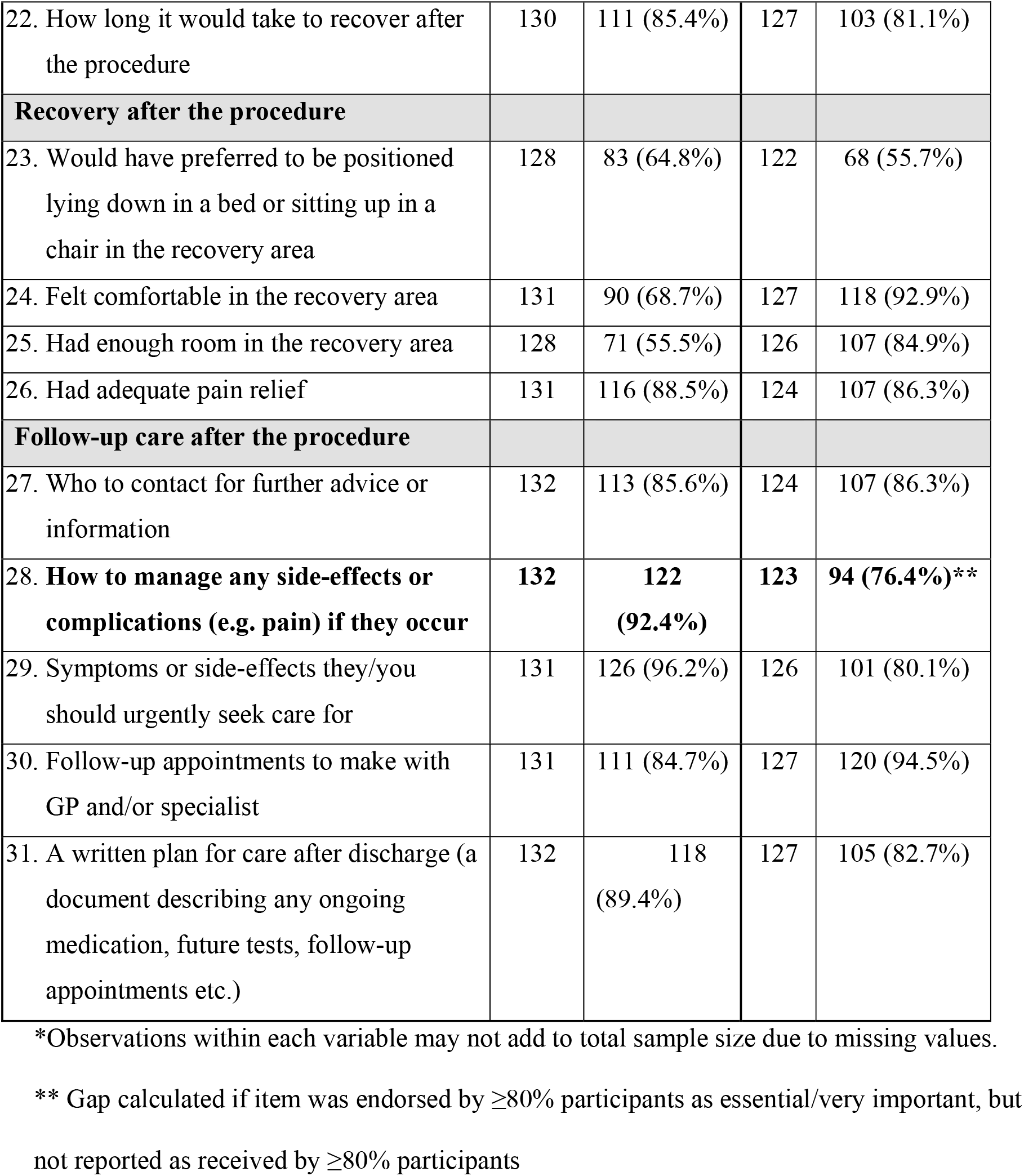
**Patients’ ratings of importance and whether items/components of care were received in relation to their cardiac catheterisation** (items identified as a gap in patient-centred care are bolded**)

### Item endorsement and identified gaps in care

Overall, most of these items (n= 43/65) were endorsed as “essential” or “very important” by 80% or more of respondents. The number of responses is reported for each individual item in Table 2. Figure 1 presents a visual summary of the total number of items examined; the number of items rated as essential/ very important by ≥80% of respondents; and the number of those for which <80% of respondents reported them as being received. Of the 43 items endorsed as essential or very important, the three items endorsed with the highest percentages for patient-reported as received were: 1) What the procedure involves (e.g. what will happen during the procedure) (92% endorsed as essential, 90% reported as received); 2) Follow-up appointments to make with their GP and/or specialist (86% endorsed as essential, 99% reported as received); and 3) Following treatment recommendations (e.g. taking medications) (87% endorsed as essential, 93% reported as received). Missing data ranged from 5-9 responses per item and were treated as missing.

**Figure 1:**
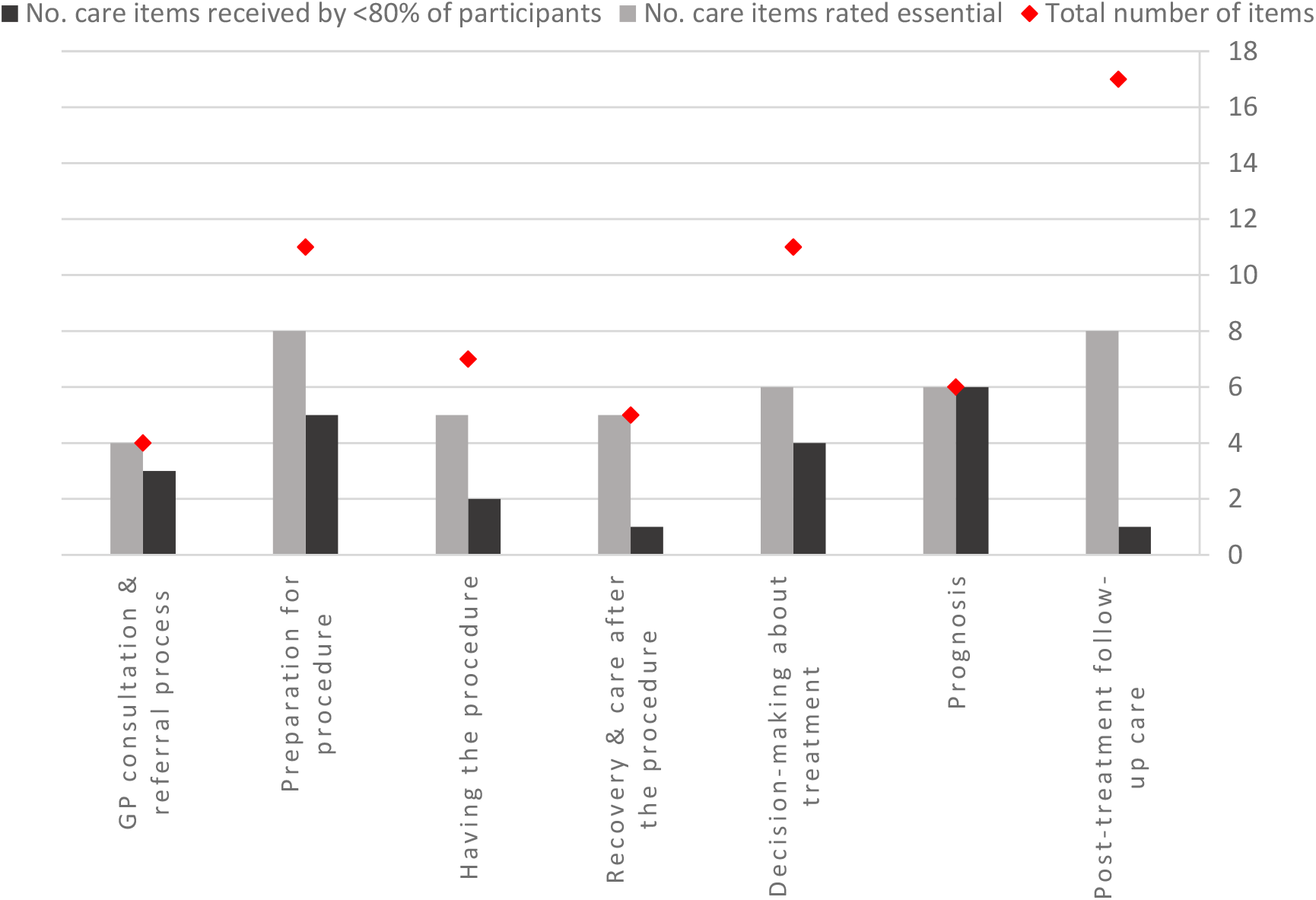
Number of items rated as essential compared with number of items reported as received by patients.

#### GP consultation and cardiologist referral process

All four items that examined this phase of care were endorsed by 80% or more of respondents as being essential or very important, with endorsement ranging from 93.9-96.9%. 82.6% of participants were told why they were being referred to a cardiologist. However, gaps in patient-centred care for receipt of the component of care were identified for the three remaining items: how to manage symptoms while waiting for their specialist appointment, what the cardiac catheterisation procedure might show, and the likely next steps after the procedure; with all having less than 80% of respondents reporting the care experience as received, with 27%, 20.3% and 23.3% of participants reporting they did not receive the item, respectively.

#### Preparation for cardiac catheterisation

Eight of the eleven items were endorsed as being essential or very important. Of the eight items, the most reported items for care received were: the expected benefits, and possible risks or complications, both highly endorsed by over 95% of respondents and received by 84.9% of participants. Gaps in the receipt of the component of care were identified for the four remaining items. However, for three items identified as having a patient-centred care gap: what could happen if the patient didn’t have the cardiac catheterisation procedure, being given information in the amount of detail wanted, and being able to talk about their fears or worries with the healthcare team; the ratings were just under the 80% threshold at 72.9%, 78.3% and 79.3% respectively.

### Having the procedure

Five of the seven items that examined this phase of care were endorsed by 80% or more of respondents as being essential or very important. What needs to happen before the procedure and what the procedure involves were both endorsed as essential and reported as experienced by 87.4% and 89.8% of participants respectively. Two gaps in patient-centred care were identified in relation to what might be experienced during the procedure, and how they might feel after the procedure, reported by 72.8% and 78.7% of participants respectively.

### Recovery after the procedure

One of the four items (adequate pain relief) was endorsed as being essential or very important. No gap in patient-centred care were identified in relation to recovery in the hospital was identified.

### Follow-up care after the procedure

All five items that examined this phase of care were endorsed by 80% or more of respondents as being essential or very important, with endorsement ranging from 84.7%-96.2%. One gap in patient-centred care was identified in relation to how to manage any side-effects or complications if they occur, reported by 76.4% of participants as received.

### Experiences across the care trajectory: subsequent cardiovascular procedures

To examine care across the whole patient care trajectory from GP referral to diagnosis and treatment inclusive of any subsequent cardiac procedures, all participants were asked whether they had undergone any subsequent cardiovascular procedures after their initial cardiac catheterisation. One hundred and seventy five respondents indicated they had undergone a subsequent related procedure including: angioplasty on a different day as the cardiac catheterisation (n=46 (17.8%)); angioplasty on the same day as the cardiac catheterisation (n=29 (11.2%)); coronary artery bypass grafting (heart bypass surgery) (n=25 (9.7%), heart valve repair or replacement (n=24 (9.3%); pacemaker insertion (n=9 (3.5%)); and cardiac defibrillator implant (n=4 (1.5%)). An additional 38 respondents (13.5%) completed the additional module of questions but did not specify which subsequent procedure they had undergone. If participants selected any of the additional procedures, they were asked to complete an additional survey module regarding care in relation to that subsequent procedure.

### Patients’ ratings of importance and whether components of care were received in relation to their subsequent cardiovascular procedure

As shown in Table 3, 130 participants completed 34 additional items identifying either: i) aspects of care that patients perceive are important or essential for a healthcare team to provide in order to best support patients receiving care in relation to their subsequent cardiovascular procedure; or ii) their actual care received in relation to: decision making about treatment (11 items); prognosis (6 items); and post-treatment follow-up care (17 items).

**Table 3.** **Patients’ ratings of importance and whether items/components of care were received in relation to their *subsequent* cardiovascular procedure** (items identified as a gap in patient-centred care are bolded**)

|  | **Important Care (n=63)**<br>*Endorsed “essential” or “very important”* |  | **Actual Care Received (n=73)**<br>**“yes” responses** |  |
| --- | --- | --- | --- | --- |
|  | *n\** | *(n, %)* | *n\** | *n (%; 95% CI)* |
| **Decision-making about treatment** |  |  |  |  |
| 1. Possible causes of the heart condition | 61 | 45 (73.8%) | 68 | 54 (79.4%, 67.8-87.6) |
| 2. Expected benefits of each treatment option | 60 | 52 (86.7%) | 69 | 59 (85.5%, 74.8-92.1) |
| 3. **Possible risks or complications of each treatment option** | **62** | **60 (96.8%)** | **69** | **53 (76.8%, 65.1-85.5)\*\*** |
| 4. **What might happen if they didn’t have any treatment** | **61** | **59 (96.7%)** | **67** | **53 (79.1%, 67.4-87.4)\*\*** |
| 5. Any ‘out-of-pocket’ costs, such as travel or co-payments for treatment | 62 | 41 (66.1%) | 65 | 27 (41.5%, 30.0-54.1) |
| 6. How long they would have to wait for treatment | 62 | 48 (77.4%) | 70 | 52 (74.3%, 62.5-83.3) |
| 7. **How involved they wanted to be in treatment decisions** | **61** | **50 (82.0%)** | **69** | **37 (53.6%, 41.6-65.3)\*\*** |
| 8. How involved they wanted their partner, family or friends in treatment decisions | 60 | 37 (61.7%) | 70 | 31 (44.3%, 32.9-56.3) |
| 9. Understanding of the goal/ purpose of treatment | 61 | 49 (80.3%) | 73 | 61 (83.6%, 72.9-90.5) |
| 10. Understanding of how long it may take to recover from treatment | 61 | 52 (85.2%) | 72 | 55 (76.4%, 65.0-85.0) |
| 11. If wanted time to think about options | 61 | 35 (57.4%) | 69 | 34 (49.3%, 37.4-61.2) |
| **Prognosis** |  |  |  |  |
| 12. Whether heart condition can be cured | 60 | 56<br>(93.3%) | 69 | 41 (59.4%, 47.2-70.6)** |
| 13. If heart condition is likely to affect life expectancy, and if so, how | 61 | 57<br>(93.4%) | 66 | 30 (45.5%, 33.6-57.8)** |
| 14. Experiencing another heart problem/condition within the next 12 months | 62 | 56<br>(90.3%) | 72 | 24 (33.3%, 23.2-45.2)** |
| 15. Needing another type of treatment in the future for the heart condition | 61 | 55<br>(90.2%) | 73 | 30 (41.1%, 30.2-52.9)** |
| 16. Being admitted to hospital within the next 12 months due to the heart condition | 61 | 54<br>(88.5%) | 72 | 21 (29.2%, 19.7-40.9)** |
| 17. Dying within the next 12 months due to the heart condition | 61 | 54<br>(88.5%) | 72 | 10 (13.9%, 7.5-24.2)** |
| **Post-treatment follow-up care** |  |  |  |  |
| 18. Managing chest pain or discomfort | 62 | 57<br>(91.9%) | 69 | 57 (82.6%, 71.5-90.0) |
| 19. Symptoms or side effects they should urgently seek care for | 61 | 60 (98.4%) | 72 | 63 (87.5%, 77.3-93.5) |
| 20. Follow-up appointments to make with their GP and/or specialist | 63 | 54 (85.7%) | 72 | 71 (98.6%, 90.4-99.8) |
| 21. When they should be able to return to work (if applicable) | 58 | 35 (60.3%) | 56 | 40 (71.4%, 57.9-82.0) |
| 22. When they should be able to return to driving | 59 | 41 (69.5%) | 60 | 54 (90.0%, 79.1-95.5) |
| 23. When they should be able to return to your usual activities (e.g. shopping, cleaning etc.) | 63 | 40 (63.5%) | 70 | 57 (81.4%, 70.3-89.0) |
| 24. Additional trustworthy information resources about their heart condition and treatment (e.g. Heart Foundation, internet sources, brochures etc.) | 62 | 38 (61.3%) | 71 | 51 (71.8%, 60.0-81.2) |
| 25. Who to contact if they have any questions or concerns | 63 | 51 (81.0%) | 73 | 63 (86.3%, 76.1-92.6) |
| 26. How to access support services and support groups within your community | 62 | 39 (62.9%) | 70 | 46 (65.7%, 53.6-76.1) |
| 27. Following treatment recommendations (e.g. taking medications) | 62 | 54 (87.1%) | 73 | 68 (93.2% 84.3-97.2) |
| 28. How to manage their mood and emotions | 63 | 43 (68.3%) | 67 | 20 (29.9%, 19.9-42.1) |
| 29. A written plan for care after discharge (a document describing any ongoing medication, future tests, follow-up appointments etc.) | 63 | 59 (93.7%) | 72 | 58 (80.6%, 69.5-88.3) |
| 30. Smoking (e.g. help to quit) | 59 | 46 (78.0%) | 56 | 23 (41.1%, 28.7-54.7) |
| 31. Alcohol (e.g. help to reduce intake) | 59 | 43 (72.9%) | 56 | 24 (42.9%, 30.3-56.4) |
| **32. Diet and nutrition** | **63** | **54 (85.7%)** | **68** | **45 (66.2%, 53.9-76.6)\*\*** |
| 33. Exercise | 62 | 53 (85.5%) | 70 | 56 (80.0%, 68.7-87.9) |
| 34. Feelings of distress, worry or sadness | 63 | 50 (79.4%) | 68 | 27 (39.7%, 28.6-52.0) |

\*Observations within each variable may not add to total sample size due to missing values.
\*\* Gap calculated if item was endorsed by $\geq 80\%$ participants as essential/very important, but not reported as received by $\geq 80\%$ participants

Overall, most of these items (n= 20) were endorsed as essential or very important by 80% or more of respondents. Similar to the items examining patients’ ratings and experiences of care related to their cardiac catheterisation, for the items that were endorsed as very important or essential by 80% or more of respondents, a high proportion of respondents also reported that these components of care were received in relation to their subsequent heart procedure. Gaps between some aspects of patient-rated important care, and the actual care received by patients, are outlined below.

### Decision-making about treatment

Six of the 11 items that examined this phase of care were endorsed by 80% or more of respondents as being essential or very important. Four gaps in patient-centred care were identified, including one item: being asked how involved they want to be in treatment decisions, which was endorsed by 82.0% of participants, but only reported as received by 53.6% of participants. In relation to the three items with a patient-centred care gap: possible risks or complications of each treatment decision, what might happen if they didn’t have treatment and understanding how long it may take to recover from treatment; the ratings were just under the 80% threshold at 76.8%, 79.1% and 76.4%, respectively.

### Prognosis

All six items that examined this phase of care were endorsed by 80% or more of respondents as being essential or very important, with endorsement ranging from 88.5%-94.4%. A patient-centred care gap was identified in relation to all six items, with the percentage of patients indicating they have received the item of care or information ranging from 13.9% in relation to whether they were explained their chance of dying within the next 12 months due to their heart condition; to being ask if they wanted to talk about whether their heart condition can be cured (59.4%).

### Post-treatment follow-up care

Seventeen items examined post-treatment follow-up care and eight were endorsed by >80% of participants as essential or very important. Only one component of care was identified as having a gap in patient-centre care: being asked about and offered help or referral if needed for diet and nutrition advice.

## DISCUSSION

This is the first Australian study and one of few internationally to examine patient-centred care among outpatients who have undergone cardiac catheterisation, one of the most widely performed cardiac procedures. Internationally, some studies have examined aspects of patient-centred care via satisfaction surveys,(21) and qualitative interviews.(22, 23) However, this is the first study to quantitatively examine patient views on what characterises essential or important care in relation to cardiac catheterisation and combine this information with patient-reported experiences to identify gaps in care.

This study also aimed to bridge another research gap by examining all preparatory content areas (risk communication, procedural information, sensory information, behavioural instruction and psychosocial aspects),(14) and care across the entire treatment pathway, from GP consultation, cardiologist referral, preparation for the procedure, diagnosis, involvement in decision-making about treatments, and follow-up care. Overall, aspects of follow-up care were highly endorsed by patients as essential components of care and well addressed, having been reported as experienced by the majority of patients. However, the findings of this study suggest that there are some gaps in patient-centred care across the care trajectory for patients.

Overall, most of the items examined (n= 43/65) were endorsed as “very important” or “essential” by 80% or more of respondents. Of those, for 22 items, less than 80% of respondents reported the care as received, indicating a gap in patient-centred care. Gaps were identified in relation to GP consultation (3 items), preparation (4 items), having the procedure (cardiac catheterisation) (2 items), follow-up care (1 item), decision making for treatment (4 items), prognosis (6 items) and post-treatment follow-up (1 item). Overall, these results suggests that outpatients may benefit from increased information provision prior to their procedure, and also specific post-procedure information in relation to their prognosis.

In particular, GP consultation and referral process and prognosis information warrant further examination. Whilst all four items that examined the GP consultation and referral process were rated as very important or essential by respondents, gaps in care were identified in relation to three of the items. However, it is recognised that for two items examined, GPs would not be considered the experts to know what the cardiac catheterisation procedure may show and what the next steps may be. There may be an expectation from the GP that the cardiologist performing or ordering the test will explain these aspects. No other quantitative studies have been identified that have examined GP information provision at the point of referral for cardiology. However, whilst not specific to cardiac catheterisation, it has been reported that “Communication between primary care physicians and specialists regarding referrals and consultations is often inadequate, with negative consequences for patients.”(24) Other research has also confirmed that patients have limited understanding of the procedure. For example, in a German cohort study of 200 patients prior to elective coronary angiography, whilst 95% of patients reported they had been well informed, less than half of the potential complications could be remembered by the patients.(25) Further studies may be appropriate to address this current clinical deficit. Development of tailored written patient education resources for the primary care environment may offer a useful adjunct to patient care; alternatively, more detailed procedural information could be provided following specialist consultation. It should be noted not every cardiology referral will result in cardiac catheterisation, which may influence the nature of the discussion between the GP and patient.

The providing of prognostic information is a difficult topic to discuss with patients given the variable natural history of cardiovascular disease and difficulty in generalising findings from population studies to individuals.(26). Furthermore, this study examined stable outpatients, so our results should be interpreted taking this into consideration. In this study, all six items that examined prognosis were endorsed by 80% or more of respondents as being essential or very important, with high endorsement ranging from 88.5%-94.4%. These findings highlight the importance patients place on this information. A patient-centred care gap was identified in relation to all six items, with the percentage of patients indicating they received the item/information ranging from 13.9% to 59.4%. Whilst uncertainty of prognosis is acknowledged, this study indicates that patients could benefit from acknowledgement and discussion of the uncertainty in relation to these topics. The opportunity to address prognosis may also be limited within the procedural setting. It is also important to acknowledge that patient may also see more than one cardiologist, and thus from whom the patient receives any such information may differ. For example, the performing cardiologist may undertake the procedure and refer the patient back to another cardiologist who may be more likely to address patient information needs based on their ongoing therapeutic relationship. In our study, for the prognosis items we asked patients if their “healthcare team explained the chance of… [item]”. Future research could investigate from whom patients may expect or prefer this information to be received. Similar research has been performed in oncology, where additional work has been done in relation to methods for breaking bad news and discussing uncertainty regarding prognosis to patients.(27)

There are few other published studies on patient experiences of cardiac catheterisation. Existing research has instead focused on patient experiences of pain and discomfort during the procedure,(28) or has utilised other approaches, such as the Lean 6 Sigma approach to quality improvement.(29) Such quality improvement work has commonly focussed on on-time patient and physician arrival, start time(30) and other clinical, organisational and procedural outcomes, including time-to-needle and other service-related factors, such as recovery statistics and MACE outcomes. Further research and publications are required to advance the field of patient experiences related to cardiac catheterisation. One study conducted in Germany examined determinants of patient satisfaction after hospitalization for cardiac catheterisation.(21) However, that study examined satisfaction, rather than patient experiences, with Likert scale response options from “excellent” to “very poor”. Thus, responses were evaluative, rather than examining actual experiences of care as in our study. In their study, the lowest ratings were reported for discharge procedures and instructions; patients were most satisfied with the kindness shown by medical practitioners and nurses. These findings similarly highlight the importance of the communication practices of healthcare professionals, with our study highlighting a particular need for caregivers to provide early procedural information and to also ensure detailed prognostic information is provided prior to discharge.

No other study has examined the cardiac catheterisation experience from the point of cardiologist referral to any subsequent cardiovascular procedures. Recent efforts to systematically measure data on patients’ perceptions of care quality have been implemented in several countries, including the USA and Australia. The suite of Picker Institute surveys are commonly used to assess patients’ experiences of care across eight patient-centred care domains.(31) However, whilst such surveys are comprehensive, they do not reflect each sequence of the care pathway for patients undergoing cardiac catheterisation, from referral through to treatment and follow-up care. Such nationwide surveys are also reported to not provide information at a local level suited for quality improvement. One of the advantages of this study was the ability to measure both patient importance, and experiences of care.

Measuring components of care across the care pathway enables areas for improvement to be identified, and healthcare providers can use this format as quality assurance tool to identify areas for improvement, and areas of excellence. For example, future work could repeat these surveys at regular intervals as part of benchmarking and continuous quality improvement initiatives.

## Limitations

Sampling bias, due to recruitment from only one public outpatient Cardiac Catheterisation Laboratory may limit the ability to generalize these findings to a broader population of cardiac catheterisation patients. This study achieved a response rate of 45%; participation necessitated completion of a pen-and-paper survey and return via mail, which may have affected participant willingness to complete. However, survey completion rates of 35-40% are commonly reported for health research and considered acceptable for routine healthcare monitoring.(32) The study measures were specifically designed for this research, and thus whilst content validity, considered the most important measurement property,(19) has been established, examination of other psychometric properties should be considered as part of future work. In addition, the two surveys (the Important Care Survey and Actual Care Survey) were sent sequentially and completed by different samples of patients, whereby patients who had undergone cardiac catheterisation 3-6 months ago answered the Important Care items, while patients who had undergone cardiac catheterisation in the last 3 months answered the Actual Care items. This may have introduced some differences in perceptions and recall bias between the two different surveys, which need to be considered when interpreting the study results.

## Conclusions

This study, designed to better understand how we can support patients before and after having cardiac catheterisation and subsequent cardiovascular procedures, has provided new knowledge regarding patient experiences of cardiac catheterisation. This study is novel in exploring all points in the cardiac catheterisation pathway from the patient perspective to identify areas where care may be improved. This research highlights how care can be improved for future patients and the information gained in this study will be used to help develop ways to improve patient care and support both before and after cardiac procedures. The findings from this study suggest ways to address gaps across the care trajectory, particularly the GP consultation and referral process and prognosis information warrant investigation.

## Data Availability

Materials for this study may be available by emailing the corresponding author. The data are not publicly available due to ethical restrictions. Additional use of or access to the data requires that the research team submit a request for variation of ethics approval.

## Acknowledgments

The authors wish to thank all participants involved in the study for their significant contribution.

## Sources of Funding

This project was supported by funding from the Priority Research Centre for Health Behaviour, University of Newcastle; and infrastructure funding from the Hunter Medical Research Institute. AL Sverdlov is supported by the Future Leader Fellowship from the National Heart Foundation of Australia (Award ID 106025).

## Disclosures

None.

## Notes

### Competing Interest Statement

The authors have declared no competing interest.

### Clinical Trial

N/A. This study as not a clinical trial.

### Author Declarations

This research was approved by the Hunter New England Human Research Ethics Committee.

